# Facilitators and barriers to contraception access and use for Hispanic American adolescent women: An integrative literature review

**DOI:** 10.1101/2024.04.10.24305616

**Authors:** Lindsay M. Batek, Natalie M. Leblanc, Amina P. Alio, Karen F. Stein, James M. McMahon

## Abstract

**Statement of the Problem:** Hispanic American adolescents experience high rates of pregnancy with profound consequences. Compared with White teens, Hispanic teens use contraception less frequently and often choose less effective forms of contraception. Access to contraception is a primary barrier to use which contributes to relatively high and disparate rates of pregnancy in Hispanic teens. This integrative review identifies facilitators and barriers of contraception access and use for Hispanic women, 13-17 years of age, living in the U.S.

**Methodology:** Following the Whittemore and Knafl (2005) methodology and PRISMA guidelines, peer reviewed studies were retrieved from PUBMED, CINAHL and EMBASE. The *Mixed Methods Appraisal Tool* was used to assess the study quality and thematic analysis was used to categorize barriers and facilitators.

**Findings:** Of 131 studies retrieved, 16 met inclusion criteria. Individual, interpersonal and healthcare provider factors were identified as primary categories of barriers and facilitators with contextual issues comprising an additional barrier category. Individual level barriers were disproportionately represented and further categorized into themes: beliefs/misconceptions, dislike of contraception, pregnancy risk perception, lack of knowledge, and lack of control. The most frequently reported facilitators were perceived parent comfort discussing sexual health and past experience of pregnancy.

**Discussion:** Limitations in this review may stem from heterogeneity in the acculturation and geography of participants and analysis by a single reviewer. Implications include considering the range of information sources and the influence they have on risk perception and risk mitigation for this population.

**Conclusion & Significance:** Adolescents describe many modifiable influences on contraception access and use. Misperceptions related to contraception, stemming from beliefs and perceptions, can be corrected through increased access to reliable sources of sex education, parental support, and direct access to nurses and healthcare providers. Awareness of these influences can inform further research and intervention development to address these health disparities.

## Introduction

Unintended teen pregnancy in Hispanic women is a significant public health problem that has profound consequences. Compared with White teens, Hispanic teens use less contraception and methods used are less effective [1, 2]. Further, Hispanic teens are less likely to have used contraception at last intercourse (24% and 12%, respectively) and to have used a prescription (more effective) method of contraception (38% for White and 18% for Hispanic teens)[3].

Hispanic teens have described significant barriers to accessing effective contraception, which contributes to relatively high rates of teen pregnancy [1]. Hispanic teen women experience teen pregnancy at a rate of 25.3 births per 1,000 women, compared with 11.4 birth per 1,000 Non-Hispanic White women [4]. In addition to being a costly problem with estimated annual healthcare costs of $6 billion in lost tax revenue and $3 billion in public expenses [5], it has significant short- and long-term consequences for teen mothers and their children [6, 7]. Teen pregnancy can affect educational attainment, economic status, and mental, social, and physical health for mother and child. The school dropout rate is higher for pregnant teens, which disrupts education and can subsequently limit employment possibilities, earning potential and increase risk of poverty [5]. Teen mothers have an increased risk of depression, substance abuse, and criminal conviction [5]. Children of teen mothers may have adverse health and developmental outcomes including prematurity, low birth weight, higher risk of perinatal and infant mortality, and poorer long term cognitive development [5]. These consequences affect long-term health trajectories and disproportionately affect Hispanic adolescents and their children [8].

### Disparities in Contraception Use and Access

According to the CDC, 75% of pregnancies in women between the ages of 15-19 years are unintended [9]. Unintended teen pregnancy is avoidable through abstinence or consistent use of effective contraception [10]. Contraceptive misuse or nonuse [11] or use of less effective contraception [12] may be partially explained by lack of knowledge about pregnancy risk, birth control, barriers to access, or distrust of the healthcare system [12–14]. The most effective methods of contraception for sexually active teens, such as oral contraceptive pills, intrauterine devices, and contraceptive implants, require a prescription [10, 13].

Healthcare access is defined as “the timely use of personal health services to achieve the best health outcomes” [15]. The Institute of Medicine definition of ‘access’ combines it with healthcare use, which can be considered “realised access” [16]. Identified barriers to contraception access include affordability, shame, embarrassment and physical difficulty reaching services and these barriers disproportionately effect individuals from marginalized populations based on age, income, race/ethnicity, rurality, and education [17]. In contrast, expanded access to family planning services has been shown to increase use of contraception and reduce teen pregnancy rates [10, 13]. However, adolescents may have little experience and fewer resources to access care and are less likely to receive adequate care when they receive services [12, 18].

Despite the myriad of known barriers to contraception access, three out of four teens are successfully finding and using contraception [1] which raises questions about the factors that determine contraception access and use and how some women are able to access contraception while others are not.

#### Definitions

This integrative review was conducted to identify literature related to the access and use of contraception among female Hispanic adolescents or teens, living in the U.S. The World Health Organization defines ‘adolescence’ within the age range of 10-19 years [19]; however, for this study we used a narrower age range of 13-17 years old because of the high rates of inconsistent contraceptive use and unintended pregnancy in this group [20]. ‘Hispanic’ is defined as “a person of Cuban, Mexican, Puerto Rican, Cuban, South or Central American, or other Spanish culture or origin, regardless of race” [21]. Given that Hispanic is an ethnic identity, individuals who identify as Hispanic may pertain to several distinct racial groups, the majority of whom identify as multiracial [22]. ‘Contraception’ is defined as the “intentional prevention of conception through the use of various devices, sexual practices, chemicals, drugs, or surgical procedures” [23]. While abstinence is included in the definition of contraception, for this review, our interest is in sexually active teens accessing contraception.

## Methods

### Study search

This integrative review follows the Whittemore and Knafl [24] methodology which is a five-step process that identifies the problem, conducts a well-defined literature search, assesses study quality, analyzes data, and articulates conclusions. The *Mixed Methods Appraisal Tool, Version 2018* (MMAT)[25], was used to assess the quality of included studies. The data from primary sources was extracted into tables and categorized, synthesized, and summarized into integrated conclusions.

A systematic search was conducted from three online databases: PUBMED, CINAHL and EMBASE. Keywords searched were (Hispanic[tiab] OR Latina [tiab]) AND (“Adolescent”[Mesh] OR “teen*”[tiab]) AND (“Contraception” [Mesh] OR “birth control”[tiab]) AND (“Access”[tiab] OR “decision” [tiab]). Additional hand searching and searching from reference lists of included studies contributed to the search.

### Eligibility Criteria

Inclusion criteria for this review were studies published in English, in peer reviewed journals, prior to February 2024 that addressed female Hispanic adolescent access to contraception or birth control. Studies were included when the sample included Hispanic adolescent women between the ages 13-17 years of age living in the United States. Studies were selected based on meeting the inclusion criteria and non-Hispanic groups, males and multiple age groups were included if data was presented separated by race/ethnicity (Hispanic), gender (female), and age (13-17 years old).

Studies were excluded if they were not peer reviewed, gray literature, non-English studies, and studies that were not related to the research questions such as intervention studies, healthcare provider strategies or studies about parental views of contraception. Intervention studies were excluded because the purpose of this review is to identify naturally occurring strategies reported by adolescents that facilitate access to contraception.

### Selection Process

Once the search was completed, duplicate studies were removed, titles and abstracts were screened for eligibility, and full texts were then reviewed for inclusion including relevance to the study aims. Excluded studies were grouped by reason for exclusion as noted on the PRISMA flow diagram (Fig 1) [26]. The initial search resulted in 131 studies, of which 16 were included in this integrative review.

**Fig 1.**
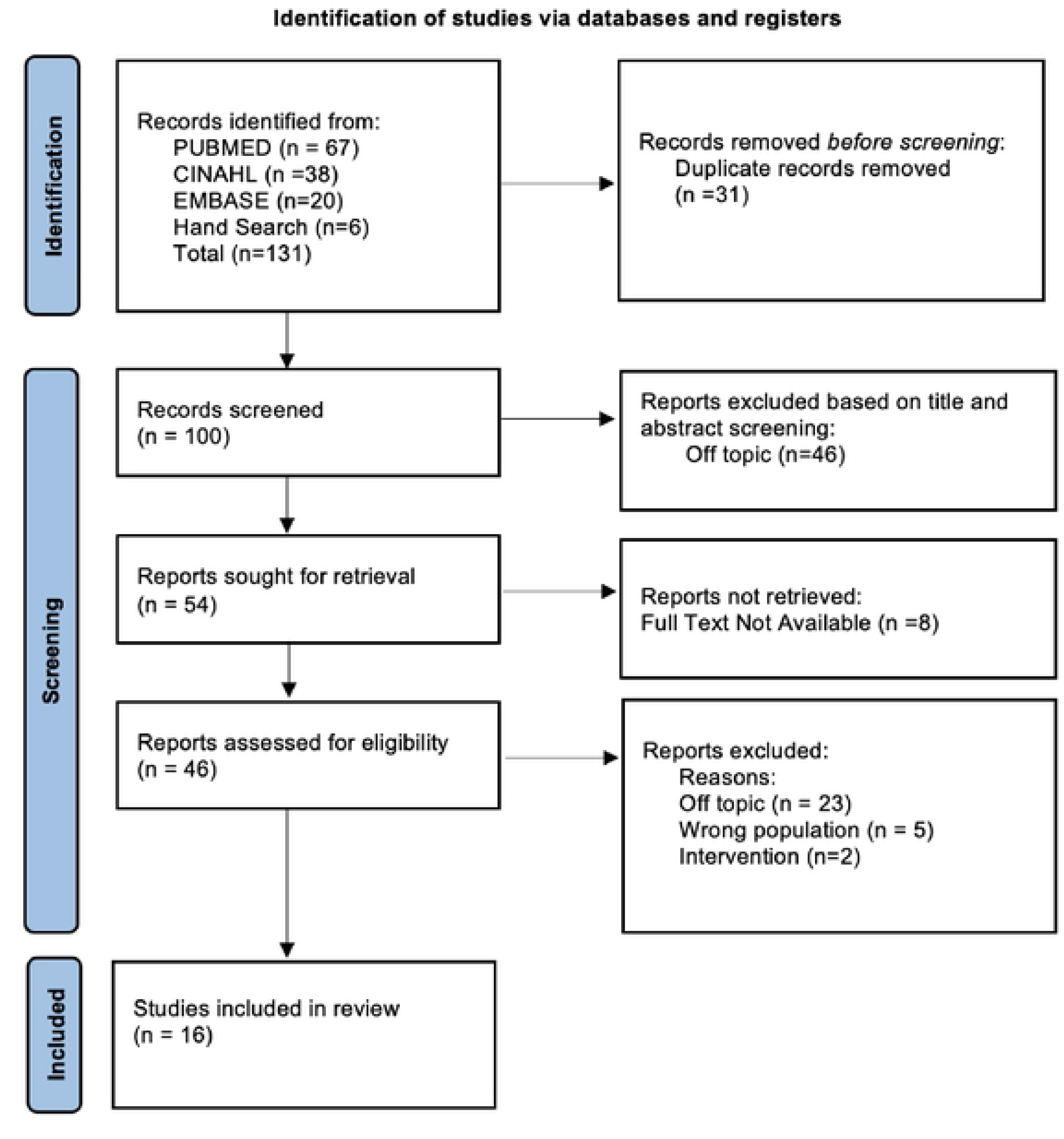
PRISMA 2020 Flow Diagram.

### Quality Appraisal

The MMAT [25] is a critical appraisal tool designed for reviews that include qualitative, quantitative, and mixed methods studies [25]. This tool facilitates the process of appraising various types of literature in one tool by answering the series of quality assessment questions as they are presented rather than calculating an overall score [25].

### Data Extraction and Synthesis

Following the process outlined by Whittemore and Knafl [24], data were extracted from the 16 selected articles including sample characteristics, location, study design, theory used, measurement tools, outcomes, barriers, and facilitators. Barriers and facilitators were identified by reviewing the data, and thematic analysis [27] was utilized to create initial codes, collate, and group codes into themes and categories. Each occurrence of a barrier or facilitator in the 16 selected studies was counted, and the total frequencies across all papers were tabulated and organized into tables and figures.

## Results

### Study Selection

Of the 131 articles identified from the database search, 31 duplicates were removed, 100 articles were screened by their title and abstract, 46 of which were excluded because they were off topic. The retrieval of full text was sought for 54 records. Full text articles were unavailable for 8 articles, and 46 were screened. Reasons for exclusion are noted on the PRISMA [26] flow diagram. The remaining 16 articles were included in the review.

#### Study methodology

This review was based on 16 peer reviewed articles published about barriers and facilitators to contraception in Hispanic adolescent women living in the U.S. This review included qualitative studies (n=11), quantitative descriptive studies (n=3), and mixed method studies (n=2). For the qualitative studies, the type of qualitative methodology (grounded theory or ethnography, etc.) was rarely described. Qualitative studies relied mostly on interviews [28–34] and focus groups [8, 11, 18, 31, 35]. Barral et al. [6] used mixed methods and grounded theory focus groups and open-ended questions. Gilliam et al. [1] used focus groups to develop a survey instrument to later give to participants. For the quantitative studies included in this review, there were very few validated scales or measurement tools that were used to measure the outcomes of interest. Of the few quantitative studies that used survey instruments, measures used included the contraceptive failure rate (CFR); contraceptive risk indices (CRI), which represents pregnancy risk; and the pregnancy risk index (PRI), which is the probability of pregnancy based on sexual activity and birth control method [2]. Two studies were secondary analyses [2, 36] and one was a retrospective chart review [13].

### Study Quality

The aim and purpose of the studies were well-aligned with the research questions, chosen methodology and interpretation of the data. The MMAT indicated that the selected studies were generally of high quality with low risk of bias although a few weaknesses were apparent, such as use of a convenience sample, conceptual issues such as grouping belief and experiences together, not explicitly stating the validity of the measures used, and a lack of information about non-responders. The results of the quality assessment are listed on the third column of Table 1.

**Table 1:** Included Studies.

| **Author details, date** | **Study objective(s)** | **Study design, Analysis, Quality Assessment** | **Sample and context** | **Authors' main conclusions** |
| --- | --- | --- | --- | --- |
| Becerra & de Anda, 1984 [28] | The perspectives of Mexican American and White adolescents were examined regarding their romantic and sexual relationships, sex education and birth control and reactions to pregnancy and motherhood. | Open-ended interviews with women who were currently pregnant, had been pregnant in the past, or had miscarried.<br>QA: Strong | 32 Mexican American and 17 White adolescents who were pregnant or had been pregnant in the past between 13-19 years old | Pregnancy was a result of not being able to view pregnancy as a potential reality for oneself. The majority of Mexican American adolescents became pregnant in the context of a long-term relationship and shared close emotional bonds with the baby's father. |
| Barral, et al., 2020 [37] | To describe the knowledge, beliefs, and attitudes about contraception among rural Latino adolescents and young adults. | Focus groups and surveys. FGs discussed attitudes, norms and perceived sexual behaviors, individual surveys assessed demographics and acculturation demographics and acculturation.<br>QA: Strong | 84 Latino youth aged 15-24 years from rural Kansas | Themes that emerged included geographical/rural location, cultural barriers, religious influences, lack of sexual education, and personal attitudes toward pregnancy and contraception use. Participants described close-knit communities with limited access to confidential reproductive health care and cultural and religious factors as influences on family planning behaviors and obstruct access to sexual health and contraception knowledge and services. Ambivalence regarding pregnancy intentions was common, along with the belief that contraception equates with abortion. |
| Carvajal et al., 2017 [35] | To identify factors that influence contraceptive decision making among Latinas in Baltimore. | Focus groups and semi structured interviews<br>QA: Strong | 8 Latina women between the ages of 15-24 years old in Baltimore, MD | The main driver of contraceptive intentions is the desire to avoid unintended pregnancy. The role of PCPs in contraceptive decision making is to build strong patient relationships through heightened communication and trust. |
| Chernick et al., 2015 [29] | To identify barriers to and enablers of contraceptive use among adolescent females using the ED and determine their interest in an ED-based pregnancy prevention intervention | Semi-structured, open ended interviews in urban ED.<br>QA: Strong | 14 predominantly Hispanic 14–19-year-old sexually active females who present to urban ED with reproductive complaints and at risk for pregnancy (nonuse of effective contraception) | Identified barriers to contraception use include side effects, mistrust in contraceptives, limited access, pregnancy ambivalence and partner pregnancy desires as barriers to hormonal contraception use. |
| Desrosiers et al., 2013[13] | To examine contraceptive choices among Hispanic and non-Hispanic girls to determine if there are differences when the barrier of cost is removed. | retrospective chart review<br>QA: Strong | 666 patients, adolescent females aged 13-19 years, 27% Hispanic | Of the 390 subjects who chose contraception during their visit, Hispanic subjects, who represented 32% of the group, were more likely to choose condoms and oral contraceptive pills compared to non-Hispanic subjects. |
| Erickson, 1998 [30] | To investigate cultural and social factors affecting the initiation of sexual intercourse among Latina adolescent mothers (who gave birth before age 18 in Los Angeles). | Life history interviews with 40 young mothers and their partners.<br>QA: Strong | 40 young, low income, Hispanic women who had births before age of 18 and their partners (only 14) conducted in 1994-1997 | The interviews suggest that for these young mothers, it was not really sex that was being negotiated, but the couple's entire relationship. "Rational" decision making regarding sex, contraception, and STD prevention could only become the norm for these young couples after they had been having intercourse for some time. |
| Galloway et al., 2017 [18] | To understand African American and Latino teens' preferences for finding health information, perceptions of accessing reproductive health services and beliefs and contraception. | 8 Focus Groups in the Fall of 2012 with African American and Latino male and female youth in South Carolina<br>QA: Strong | A total of 63 participants, African American and Hispanic youth age 15-19, participated in focus groups divided by gender and race/ethnicity | Teen concerns included confidentiality of reproductive health services, a desire for services that are private, friendly and welcoming. They also revealed inaccurate beliefs about the teens ability to consent to care without a parent's permission and about the use and effectiveness of birth control. |
| Gilliam et al., (2004) [11] | To understand factors influencing use and nonuse of contraception in young, low-income Latina adolescents through focus groups | Focus Group discussions with Latino females<br>QA: Strong | 40 Latina women between the ages of 18-26 who were recruited from an outpatient clinic in Chicago | There was complete consensus throughout the sessions regarding cultural and social influences prohibiting sex outside of marriage, admonishments against premarital sex appeared to be directed against women which contribute to contraception non-use. Following pregnancy participants had greater access to contraception and more resolve to use it. |
| Gilliam et al., 2011 [1] | To understand the factors influencing the use of effective methods of contraception in Latina adolescents and young adults as well as the relative psychosocial factors. | Face to face interviews were conducted to develop a survey instrument that was later given to participants<br>QA: Strong | 274 Non-pregnant Latina, predominantly Mexican females, between 13-25 years of age on the West side of Chicago | Study findings included the importance of tailoring messages to Latina adolescent and young adults to reduce unintended pregnancy. Interventions to improve effective contraceptive use among Latina adolescents should encourage effective forms of contraception combined with communication with their partners about birth control while considering acculturation. |
| Guilamo-Ramos et al, 2019 [38] | To explore maternal and adolescent perspectives on the involvement and role of Latino mothers in providing guidance regarding contraception use. | Focus Groups with adolescent-mother dyads (semi structured guides were used in focus groups)<br>QA: Strong | 21 mother-adolescent Latino dyads (n=42) (predominantly Puerto Rican and Dominican). Latino adolescents ages were 17-19 years | Results suggest asymmetric priorities and preferences with respect to maternal involvement in contraceptive decision making among older adolescents. Results demonstrate a missed opportunity for Latino mothers to support their older adolescent children to prevent unplanned pregnancies, STIs and HIV. |
| Kramer et al., 2018 [36] | To investigate influences (demographic, socioeconomic, race/ethnicity and reproductive health characteristics) on long-acting reversible contraceptive (LARC) use | Secondary Data Analysis with Data derived from 2011-2013 and 2013-2015 National Surveys of Family Growth (NSFG), using logistic regression analysis<br>QA: Strong-moderate | 9321 Non-Hispanic White, non-Hispanic Black, and Hispanic women of reproductive age (15-45) (11% Hispanic) | Similar patterns of LARC use were found by race/ethnicity. The experience of unintended pregnancy among whites and Hispanics predicted probability of LARC use |
| Mitchell et al., 2023 [31] | To compare contraceptive access experiences of pregnant and non-pregnant Mexican-origin youth in Mexico and California. | Focus groups, in-depth interviews and brief sociodemographic survey conducted in English and Spanish were used.<br>QA: Strong | 74 Pregnant or parenting Mexican-origin female youth from Mexico (n=49) and California (n=25) age 14-20 years. | In both locations participants described obstacles in accessing preferred contraception due to social, cultural and institutional dynamics. Participants worried about parental and peer opinions as well as side effects. Lack of knowledge about options was found in California participants. |
| Morales-Alemán et al., 2020 [32] | To examine young Latina women's perceptions and experiences of accessing sexual healthcare | Semi-structured qualitative interviews<br>QA: Strong-moderate | 20 young Latina women between the ages of 15-19 living in Alabama and in the US for $\geq 5$ years | Describe influences on accessing sexual healthcare based on the Socioecological Model of Sexual Health. Participants described barriers related to sociocultural influences including need for interpreters, transportation, feeling stereotyped; due to social relationships such as parental influences, and individual level embarrassment, stigma, or lack of awareness. |
| Norris & Ford, 1992 [33] | To identify condom beliefs that may influence intentions and use in young minority populations | Qualitative, face to face interviews, structured and with open-ended questions<br>QA: moderate | 64 African American and Hispanic (majority from Puerto Rico) adolescents (ages 15-21) (15 were Hispanic females) | Only 13% of Hispanic participants knew that nonlatex condoms do not provide effective protection. Condom use was inconsistent, with many describing negative experiences with condoms including condom breakage, slipping, reducing sensation; Hispanic females reported lower "thinking about condoms", 50% thought about using condoms |
| Waddell et al., 2010 [2] | To identify risk components such as race/ethnicity, neighborhood by modeling demographic difference, odds of recent sexual activity and birth control use | Data from the 2005 and 2007 New York City Youth Risk Behavior Surveys, (32.7% Hispanic), was analyzed to calculate overall pregnancy risk as it relates to race/ethnicity, grade level, age, borough and school neighborhood.<br>QA: Strong-moderate | 6,608 non-Hispanic Black, Hispanic and non-Hispanic - White teen girls in NYC public schools in 2005 and 2007 | Hormonal contraception use was low in all groups. Compared with non-Hispanic Whites, Hispanic female students were less likely to report use of condoms, use of a hormonal method, or withdrawal method and had a 50% increased Pregnancy Risk Index. The Contraceptive Failure Rate (predicted annual number of pregnancies per 100 sexually active public high school females was significantly higher in Hispanic females compared with non-Hispanic whites |
| Wilson et al., (2011) [34] | To explore factors that influence adolescents' postpartum use of contraceptive | In-depth interviews<br>QA: Strong | In-depth interviews with 21 black, white and Latina (n=7) adolescent first-time mothers (who gave birth between 13–17-year-old) from rural and urban areas of North Carolina | Contraceptive use increased after childbirth related to change in pregnancy attitudes and intentions, knowledge and access to contraception, improved parent communication. |

### Study Characteristics

*Study Size and Geography.* The sample size in the studies ranged from small (n=8) in the qualitative studies, to large, the National Survey of Family Growth, N=9321. One study had focus groups from Mexico and California [31] and all of the remaining studies took place in the U.S. in both rural [6, 34] and urban areas [1, 11, 35]. Some studies described the state or city locations, with represented regions in the U.S. including the East Coast (Baltimore [35]); Midwest (Kansas [6], Chicago [1, 11]); the South (Alabama [32], North Carolina [34]); and the West Coast (California [31]).

#### Demographic Descriptions

Frequent labels used to describe participants included Latino or Hispanic youth [1, 2, 6, 8, 13, 18, 29, 30, 32–34, 36]. Some studies described the samples (at least partly) as Mexican American [11, 28, 31] and immigrant Latinas [35]. The countries and places of origin of participants were not always described but those reported were Mexican [28, 31, 33], Puerto Rican [8, 33], and Dominican [8].

#### Acculturation

The acculturation level of the participants was inconsistently provided. Some studies describe the acculturation level of participants in the results [1, 6, 28, 31, 33]. When acculturation was described, it was by place of birth [30, 32, 34, 35], length of time in the U.S. [32], or acculturation score which was determined by a language based scale [1, 33] or a modified version of the Short Acculturation Scale for Hispanics (SASH) combined with years living in the US [6].

#### Theoretical Frameworks

Some studies articulated a theory that guided their research such as the Theory of Planned Behavior [34, 35], the Health Belief Model [29], the Socioecological Model of Sexual Health [32], Pechansky and Thomas’ Theory of Access [31], and the Construct Accessibility Model [33].

### Facilitators and Barriers to Contraception Described by Category

Through the data extraction and synthesis process, both the facilitators and barriers were grouped into the following overarching categories: individual, interpersonal (family, spouse/partner, friend), and healthcare provider levels; further, contextual matters emerged as an additional category of barriers. Each of these main categories was further subdivided into thematic subcategories as displayed in two sunburst charts, one for facilitators (Fig 2), and one for barriers (Fig 3). On both charts, the categories that resulted from the thematic analysis are visible in concentric rings with themes in the center and related subthemes on the periphery. The sizes of the parcels in each chart are proportional to the number of occurrences of that barrier or facilitator category across the 16 selected studies.

**Fig 2.**
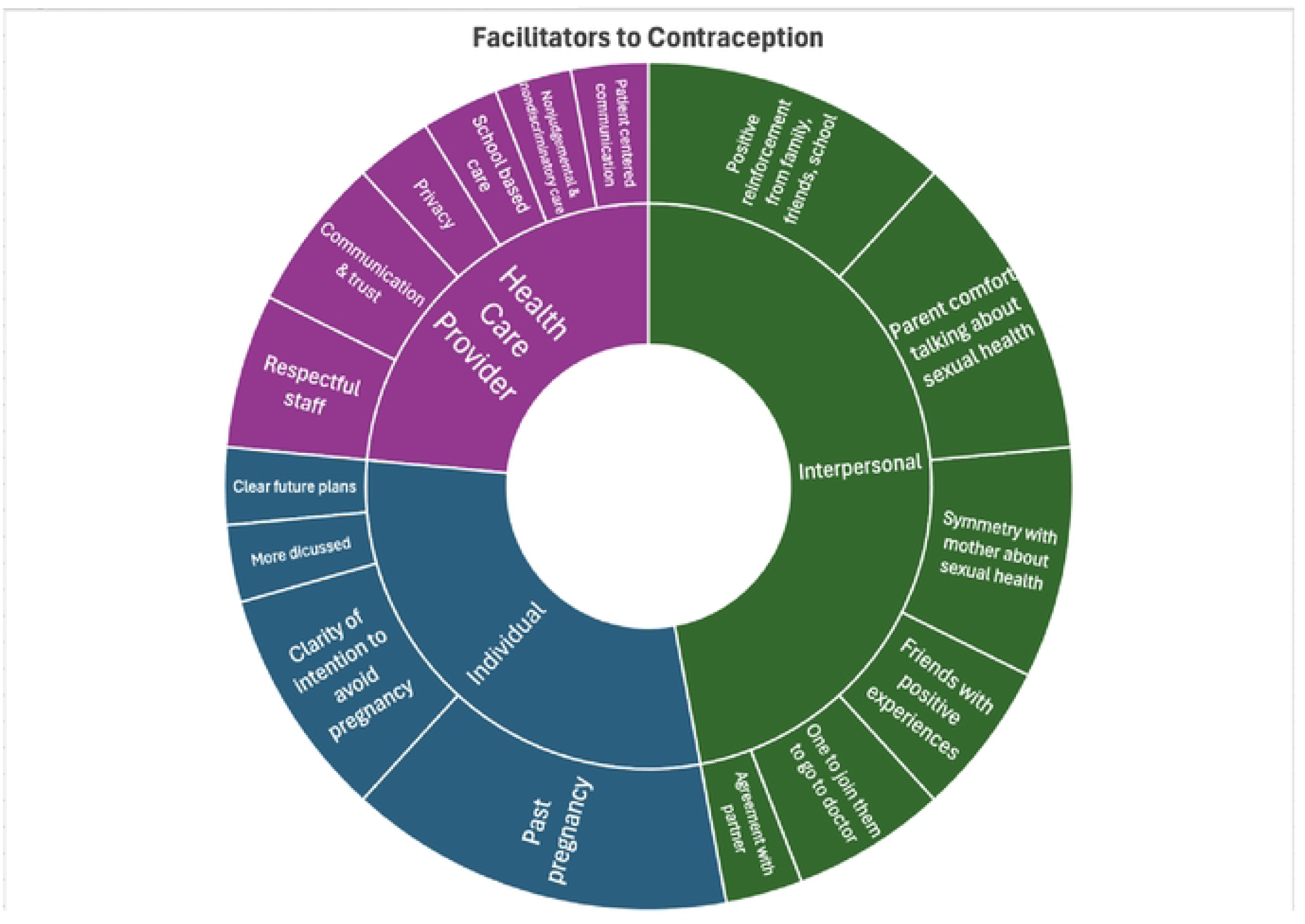
Facilitators to Contraception. Themes identified as facilitators to contraception use are displayed in the inner circle and are further differentiated into the subthemes located on the outer circle.

**Fig 3.**
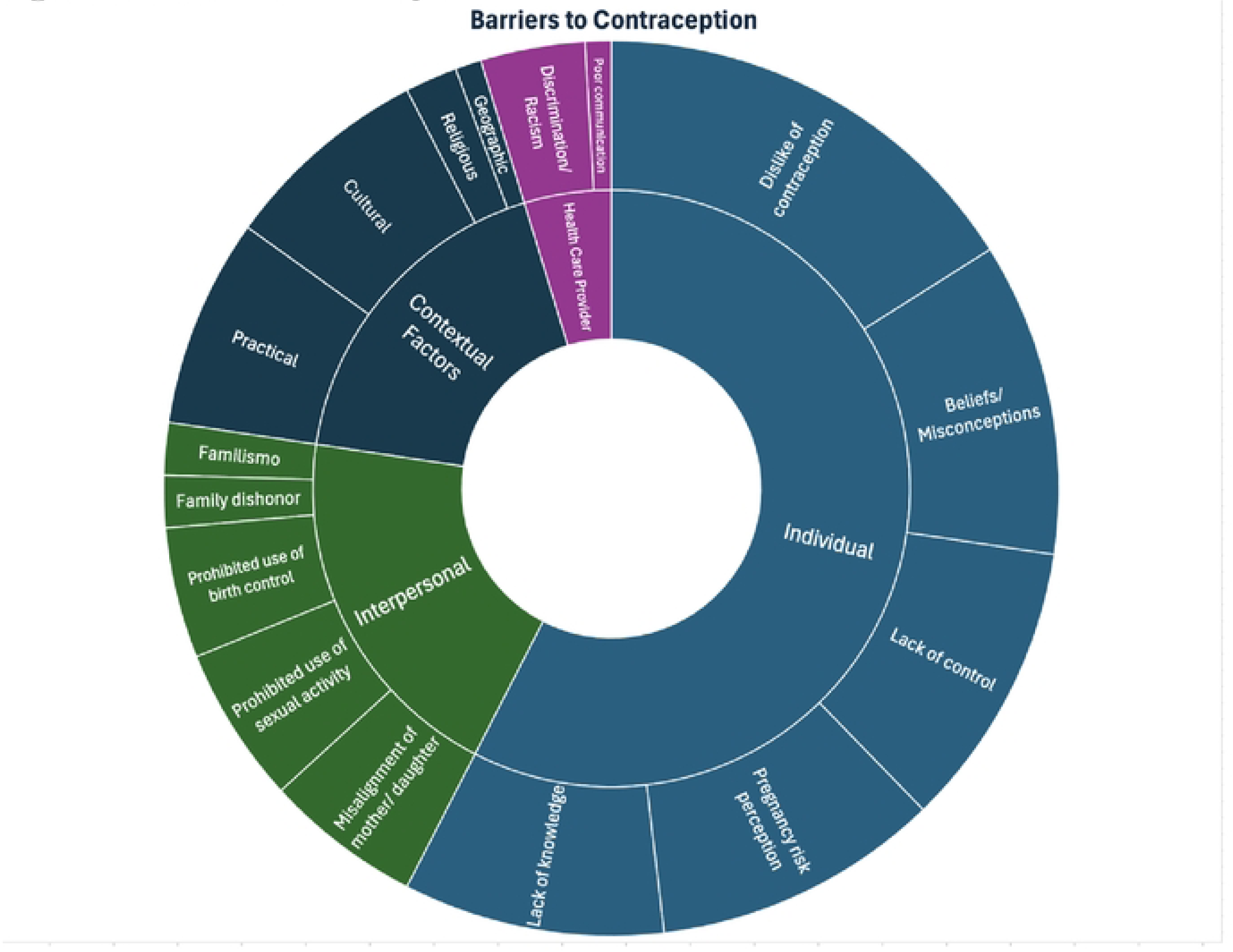
Barriers to Contraception. Themes identified as barriers to contraception use are displayed on the inner ring by with subthemes located on the outer ring.

### Category 1: Individual Level Facilitators and Barriers

The most frequently mentioned individual facilitators were the clarity of intention to avoid pregnancy [11, 34, 35], and experience of pregnancy [1, 6, 28, 34, 36]. Additional facilitators identified were clear plans for the future [29], and more communication about contraception [8, 33].

The individual level barriers constituted a disproportional amount of the total barriers identified in this review with the number of data points nearly triple that of any other category. Individual barriers to contraception access were divided into five subthemes. The subthemes include beliefs/misconceptions, dislike of contraception, pregnancy risk perception, lack of knowledge, and lack of control. Specific factors that were most frequently described as barriers were side effects/perceived health risks [11, 18, 34], misconception regarding contraception or fertility [18, 29, 32], ambiguous or ambivalent desire for children [6, 11, 28, 29], lack of knowledge about options [11, 28, 30, 33–35], and male partner dominance [11, 29, 30, 35]. Across the five subthemes, dislike of contraception and beliefs/misconceptions were the most common. The first theme, ‘dislike of contraception’ was related to the side effects such as effects on menstruation, weight and future fertility and perceived health risks of contraception [11, 18, 31, 34], that it decreases physical or emotional pleasure [18, 33], taboo [11, 30], fear of lack of confidentiality and reaction of others’ if they learn one is using contraception, uncertainty in selecting contraception [18, 29, 31], and negative past experience, such as condom breakage or slippage [29, 33].

Within the second theme, ‘beliefs/misconceptions’ there were descriptions of beliefs [18, 28], such as “abortion is murder” [6, 28], and that contraception equals abortion, that withdrawal is effective if pregnancy is not desired [11], mistrust of the healthcare system [29, 35], and misconceptions regarding contraception and fertility [18, 29, 32].

The third theme, ‘pregnancy risk perception,’ encompasses several attitudes and beliefs, including the view that there is no risk of pregnancy [18, 28, 31], avoidance of condoms, perceiving them as protection against sexually transmitted infections rather than for pregnancy prevention [33], and ambiguous or actual desire for children [6, 11, 28, 29, 34].

Lack of knowledge and lack of control were the final two subthemes in this category. Participants described a lack of information about contraceptive options [11, 28, 30–35], feelings of embarrassment, stigma or lack of confidence accessing care [32, 34], and a lack of knowledge about biology and physiology [30]. Lack of control related to male partner dominance [11, 29, 30, 35], reliance on the male partner for the withdrawal method [11], lack of control over pregnancy related to infertility beliefs, religious influence and feelings of invincibility [11, 18, 28], uncertainty about the future [29], unplanned or unexpected intercourse [30, 34], and not considering condom use [33].

### Category 2: Interpersonal Level Facilitators and Barriers

Interpersonal facilitators to contraception access included parent comfort in discussing sexual health [8, 32, 34], accompaniment and transportation to the doctor [32, 34], positive reinforcement from family, friends and schools [1, 8, 31, 35], mother/daughter alignment or parent support on sexual and reproductive health (SRH) [8, 32, 34], friends who have had positive experiences with contraception [29, 31], and mutual agreement with partners about not wanting a pregnancy [29].

Interpersonal barriers to contraception access were described as familial influences such as *familismo* (“a value that reflects individuals’ strong attachment and solidarity with their nuclear and extended families”) [6, 11], family dishonor [6, 11], cultural or familial beliefs prohibiting use of birth control [11, 18, 31, 32, 34], and sexual activity [11, 28, 30, 32], and misalignment of parent/daughter priorities [8, 11, 29, 32, 34].

Guilamo-Ramos et al. [8] and Morales-Alemán et al. [32] explored the vital role of the maternal and parent influence on the daughter’s SRH. The need for support was expressed by several teens “My mother doesn’t really understand …. If only my mom could get it and understand I could really use her support.” [8]. And “…I pretend I have never done it. That way, I can avoid a fight.” [8]. Morales-Alemán et al. [32] reported that the main barrier to accessing SRH services for most participants was parental disapproval of them having sex and using these services. While some mothers were supportive and proactive in pregnancy prevention for their teens, others were unsure, as one parent expressed, “Should I mention it? It’s really confusing and scary because there is so much I do not know. Nobody ever told me. I got pregnant early on.” [8]. These experiences reflect how the maternal influence can be either a facilitator or barrier to SRH uptake.

### Category 3: Healthcare Provider Barriers and Facilitators

Characteristics of healthcare providers that facilitated access to contraception included patient-centered communication, nonjudgmental and non-discriminating interactions, and trustworthiness [29, 35]. Clinic features that facilitate access were providing privacy [18], having respectful office staff [29, 30], and being located in a school-based health clinic [29]. Healthcare provider barriers were low patient-rated communication [35], perceived discrimination [18, 32, 35], or inadequate care given due to race [35].

### Category 4: Contextual Level Barriers to Contraception

Contextual level influences on contraception were described in studies only as barriers (no facilitators were described) to contraception and categorized into cultural, religious, geographical, and practical influences. Culturally, sexual taboo or stigma [11, 30, 32, 39], placing a high value on virginity [6, 30], and valuing sexual ignorance and sexual passivity in women [30] are all barriers to contraception. Both religious objections to the use of contraception [1, 35] and neighborhood contexts that influence younger sexual initiation [2] were contextual influences that represented barriers to contraceptive access. One study by Carvajal et al. [35] described contradictory findings regarding the influence of religious ideology and norms. Norris & Ford [33] report a specific stigma found in their study: the perception that condoms are for unclean people. Practical matters that were barriers to contraception were loss of Medicaid and continuity of care (related to the end of pregnancy) [34], lack of time for a clinic visit [32], lack of public transit or transportation [29, 32], lack of insurance and cost [11, 32, 34], and need for interpreters [32]. Additionally, two studies reported that participants mentioned the negative influence of media and internet advertisements about contraception as inducing fear [18, 29].

## Discussion

This analysis sought to better understand barriers and facilitators that influence access to and use of contraception in female Hispanic teens ages 13-17 years old. Sixteen studies were included, and 40 specific barriers and 16 facilitators were found and presented as subthemes. The facilitators that emerged from this analysis provide insight into potentially outcome-changing influences that support women’s reproductive autonomy, decision making, and acquisition of contraception.

Among barriers identified, sentiments such as “abortion is murder” or that contraception is equivalent to abortion [6, 28], and the belief that withdrawal will work if pregnancy is not desired [11] illustrates how ideology can intertwine with misconceptions leading to inaccurate perceptions about risk and risk management. These views, combined with barriers such as embarrassment, stigma and lack of confidence accessing care [32, 34] and lack of knowledge about biology and physiology [30], highlight the complex influences that make it challenging for teens to seek information and ask questions from reliable sources such as trusted adults and healthcare providers. This complexity can contribute to a sense of lack of control, exacerbated by male partner dominance (with age difference) [11, 29, 30, 35] and reliance on the male partner for the withdrawal method [11], and a lack of control over pregnancy due to beliefs about fertility, religious influences, and feelings of invincibility or the perception that ‘it won’t happen to me’ [11, 18, 28]. Interventions that inadequately address these barriers in meaningful ways may be less effective or ineffective.

In contrast, key facilitators include a clear intention to avoid pregnancy, parental comfort discussing sexual health, positive reinforcement from family, friends and school, and good communication and trust and respect from healthcare provider and their office staff. These are actionable items that warrant consideration in the development of future interventions.

## Implications

### The Role of Sex Education and Experience

When considering how an individual makes decisions regarding contraception, perception and understanding of risk and ease of mitigating risk are each important. This review demonstrated the different sources of influence ranging from individual beliefs to personal experience of pregnancy, to people who directly support and influence teens such as parents, peers, partners, and healthcare providers. Many of the barriers identified in this review are addressed in sex education classes in schools in the U.S. including misconceptions and lack of knowledge; yet, Hispanic students may receive less sex education because they are more likely to drop out of school [40] or move frequently, which may preclude them from receiving adequate sex education [6]. Even when Hispanic students receive sex education in school, the quality and content provided is inconsistent ranging from abstinence-only to comprehensive sex education leading to disparities in what is learned, and related outcomes [41]. These disparities contribute to socially driven health and systemic inequities for Hispanic teen pregnancy rates [41].

Parents, peers, partners, and healthcare providers can have a strong influence on contraception access and can serve as barriers or facilitators. Morales-Alemán’s et al. [32] finding that parent disapproval of teens having sex was their main barrier to access to care for SRH, which indicates the importance of parent attitudes towards teen access to contraception as either a barrier [11, 32, 34] or facilitator [32]. Male partners can be barriers through male partner dominance and controlling withdrawal or facilitators through mutual agreement with partner about pregnancy intentions, positive reinforcement, providing contraception or supporting access to the healthcare providers. Healthcare providers are positioned to prime parents to be supportive of teen’s access to care for SRH. They can also support teens independently by providing care that is private, confidential, nonjudgmental, non-discriminatory, and patient-centered and in an accessible location.

Further research can build upon the findings presented here by discerning what meaningfully differentiates those who have adequate awareness of pregnancy risk and the ability to access contraception from those who do not. This may reveal the relative impact of different factors on teens, how these factors synergize to serve as barriers or facilitators, and patterns in how these influences affect teen contraceptive access and use. Understanding these dynamics may identify targeted strategies to reduce disparities in teen pregnancy.

## Limitations

The studies included in this analysis grouped teens by the umbrella terms of ‘Hispanic’ or ‘Latina’ but there is considerable diversity and variation within this group. Though commonalities such as taboos, stigma, and misconceptions were found, it is unclear if more research across ethnic or national subgroups may reveal important differences. The presence of bias in the findings presented in this review cannot be ruled out as a single author [LB] conducted the literature search, assessed studies for inclusion, rated bias, extracted, analyzed, and synthesized the data. The search sought to identify studies about Hispanic teens but did not search all potential places of origin of Hispanics in the U.S. such as Mexican, Guatemalan, Puerto Rican, Columbian etc., so studies about those specific groups, while applicable, may not have been identified.

## Conclusion

This integrative review sheds light on the barriers and facilitators to contraceptive access from the perspective of female Hispanic teens, aged 13-17 in the U.S. The synthesis of perspectives and experiences revealed in this review is informative and significant, given the disparities in contraception access and pregnancy rates in this group. Identification of barriers and facilitators can stimulate discussion and research aimed at improving the delivery of education and healthcare to meet the contraceptive needs of female Hispanic teens. The implications presented here underscore the importance of addressing the unique multilevel barriers and leveraging facilitators encountered by this population to promote health equity and improve access to contraception.

## Data Availability

I am unable to share any additional pertinent data.

## Notes

### Competing Interest Statement

The authors have declared no competing interest.

### Funding Statement

The project (LB) described in this publication was supported by the University of Rochester CTSA award number UL1 TR002001 from the National Center for Advancing Translational Sciences of the National Institutes of Health. The content is solely the responsibility of the authors and does not necessarily represent the official views of the National Institutes of Health.

### Author Declarations

No IRB approval required for this integrative review article.

## References

1. Gilliam ML, Neustadt A, Whitaker A, Kozloski M. Familial, cultural and psychosocial influences of use of effective methods of contraception among Mexican-American adolescents and young adults. J Pediatr Adolesc Gynecol. 2011;24(2):79–84. Epub 20101203. doi: 10.1016/j.jpag.2010.10.002. PubMed PMID: 21126893; PubMed Central PMCID: PMCPMC5145289.

2. Waddell EN, Orr MG, Sackoff J, Santelli JS, Waddell EN, Orr MG, et al. Pregnancy risk among black, white, and Hispanic teen girls in New York City public schools. Journal of Urban Health. 2010;87(3):426–39. doi: 10.1007/s11524-010-9454-4. PubMed PMID: 105203990. Language: English. Entry Date: 20100924. Revision Date: 20200708. Publication Type: journal article.

3. Caudillo ML, Hickman SN, Simpson SS. Racial and Ethnic Differences in the Relationship Between Risk-Taking and the Effectiveness of Adolescents’ Contraceptive Use. Perspect Sex Reprod Health. 2020;52(4):253–64. Epub 20201228. doi: 10.1363/psrh.12165. PubMed PMID: 33372342; PubMed Central PMCID: PMCPMC10506860.

4. Martin JA, Hamilton BE, Osterman MJK, Driscoll AK. Births: Final Data for 2019. Natl Vital Stat Rep. 2021;70(2):1–51. PubMed PMID: 33814033.

5. Coyne CA, D’Onofrio BM. Some (but not much) progress toward understanding teenage childbearing: a review of research from the past decade. Adv Child Dev Behav. 2012;42:113–52. doi: 10.1016/b978-0-12-394388-0.00004-6. PubMed PMID: 22675905; PubMed Central PMCID: PMCPMC3654402.

6. Barral RL, Cartujano B, Perales J, Ramirez M, Cowden JD, Trent ME, et al. Knowledge, beliefs, and attitudes about contraception among rural Latino adolescents and young adults. J Rural Health. 2020;36(1):38–47. Epub 2019/08/21. doi: 10.1111/jrh.12390. PubMed PMID: 31430396.

7. Johnson R, Nshom M, Nye AM, Cohall AT. There’s always Plan B: adolescent knowledge, attitudes and intention to use emergency contraception. Contraception. 2010;81(2):128–32. Epub 2010/01/28. doi: 10.1016/j.contraception.2009.08.005. PubMed PMID: 20103450.

8. Guilamo-Ramos V, Bowman AS, Benzekri A, Ruiz Y, Beltran O. Misalignment of sexual and reproductive health priorities among older Latino adolescents and their mothers. Contraception. 2019;99(3):179–83. Epub 2018/11/25. doi: 10.1016/j.contraception.2018.11.011. PubMed PMID: 30471265.

9. Center for Disease Control and Prevention. Unintended Pregnancy. March 27, 2023.

10. Manlove J, Ryan S, Franzetta K. Patterns of contraceptive use within teenagers’ first sexual relationships. Perspect Sex Reprod Health. 2003;35(6):246–55. doi: 10.1363/psrh.35.246.03. PubMed PMID: 14744656; PubMed Central PMCID: PMCPMC1473988.

11. Gilliam ML, Warden MM, Tapia B. Young Latinas recall contraceptive use before and after pregnancy: a focus group study. J Pediatr Adolesc Gynecol. 2004;17(4):279–87. Epub 2004/08/04. doi: 10.1016/j.jpag.2004.05.003. PubMed PMID: 15288030.

12. Dehlendorf C, Park SY, Emeremni CA, Comer D, Vincett K, Borrero S. Racial/ethnic disparities in contraceptive use: variation by age and women’s reproductive experiences. Am J Obstet Gynecol. 2014;210(6):526.e1-9. Epub 20140201. doi: 10.1016/j.ajog.2014.01.037. PubMed PMID: 24495671; PubMed Central PMCID: PMCPMC4303233.

13. Desrosiers F, Arden M, Fisher M. Patterns of contraception choice among Hispanic and non-Hispanic female adolescents. Int J Adolesc Med Health. 2013;25(2):167–70. Epub 2013/01/22. doi: 10.1515/ijamh-2013-0025. PubMed PMID: 23334055.

14. Hall KS. The Health Belief Model can guide modern contraceptive behavior research and practice. J Midwifery Womens Health. 2012;57(1):74–81. Epub 20111215. doi: 10.1111/j.1542-2011.2011.00110.x. PubMed PMID: 22251916; PubMed Central PMCID: PMCPMC3790325.

15. Office of Disease Prevention and Health Promotion. Access to health services. n.d.

16. Levesque JF, Harris MF, Russell G. Patient-centred access to health care: conceptualising access at the interface of health systems and populations. Int J Equity Health. 2013;12:18. Epub 20130311. doi: 10.1186/1475-9276-12-18. PubMed PMID: 23496984; PubMed Central PMCID: PMCPMC3610159.

17. Swan LET. The impact of US policy on contraceptive access: a policy analysis. Reprod Health. 2021;18(1):235. Epub 20211122. doi: 10.1186/s12978-021-01289-3. PubMed PMID: 34809673; PubMed Central PMCID: PMCPMC8607408.

18. Galloway CT, Duffy JL, Dixon RP, Fuller TR. Exploring African-American and Latino Teens’ Perceptions of Contraception and Access to Reproductive Health Care Services. Journal of Adolescent Health. 2017;60:S57-S62. doi: 10.1016/j.jadohealth.2016.12.006. PubMed PMID: 121275111. Language: English. Entry Date: 20170222. Revision Date: 20180530. Publication Type: Article.

19. Singh JA, Siddiqi M, Parameshwar P, Chandra-Mouli V. World Health Organization Guidance on Ethical Considerations in Planning and Reviewing Research Studies on Sexual and Reproductive Health in Adolescents. J Adolesc Health. 2019;64(4):427–9. doi: 10.1016/j.jadohealth.2019.01.008. PubMed PMID: 30904091; PubMed Central PMCID: PMCPMC6496912.

20. Magnusson BM, Crandall A, Evans K. Early sexual debut and risky sex in young adults: the role of low self-control. BMC Public Health. 2019;19(1):1483. Epub 20191108. doi: 10.1186/s12889-019-7734-9. PubMed PMID: 31703650; PubMed Central PMCID: PMCPMC6839049.

21. Aragones A, Hayes SL, Chen MH, González J, Gany FM. Characterization of the Hispanic or latino population in health research: a systematic review. J Immigr Minor Health. 2014;16(3):429–39. doi: 10.1007/s10903-013-9773-0. PubMed PMID: 23315046; PubMed Central PMCID: PMCPMC4518558.

22. Lopez M, Krogstad J, Passel J. Who is Hispanic? September 5, 2023.

23. Jain R, Muralidhar S. Contraceptive methods: needs, options and utilization. J Obstet Gynaecol India. 2011;61(6):626–34. Epub 20120214. doi: 10.1007/s13224-011-0107-7. PubMed PMID: 23204678; PubMed Central PMCID: PMCPMC3307935.

24. Whittemore R, Knafl K. The integrative review: updated methodology. J Adv Nurs. 2005;52(5):546–53. doi: 10.1111/j.1365-2648.2005.03621.x. PubMed PMID: 16268861.

25. Hong QN, Gonzalez-Reyes A, Pluye P. Improving the usefulness of a tool for appraising the quality of qualitative, quantitative and mixed methods studies, the Mixed Methods Appraisal Tool (MMAT). J Eval Clin Pract. 2018;24(3):459–67. Epub 20180221. doi: 10.1111/jep.12884. PubMed PMID: 29464873.

26. Page MJ, McKenzie JE, Bossuyt PM, Boutron I, Hoffmann TC, Mulrow CD, et al. The PRISMA 2020 statement: an updated guideline for reporting systematic reviews. Syst Rev. 2021;10(1):89. Epub 20210329. doi: 10.1186/s13643-021-01626-4. PubMed PMID: 33781348; PubMed Central PMCID: PMCPMC8008539.

27. Thomas J, Harden A. Methods for the thematic synthesis of qualitative research in systematic reviews. BMC Med Res Methodol. 2008;8:45. Epub 20080710. doi: 10.1186/1471-2288-8-45. PubMed PMID: 18616818; PubMed Central PMCID: PMCPMC2478656.

28. Becerra RM, de Anda D. Pregnancy and motherhood among Mexican American adolescents. Health Soc Work. 1984;9(2):106–23. Epub 1984/01/01. doi: 10.1093/hsw/9.2.106. PubMed PMID: 6724424.

29. Chernick LS, Schnall R, Higgins T, Stockwell MS, Castaño PM, Santelli J, Dayan PS. Barriers to and enablers of contraceptive use among adolescent females and their interest in an emergency department based intervention. Contraception. 2015;91(3):217–25. Epub 2014/12/17. doi: 10.1016/j.contraception.2014.12.003. PubMed PMID: 25499588; PubMed Central PMCID: PMCPMC4352549.

30. Erickson PI. Cultural factors affecting the negotiation of first sexual intercourse among Latina adolescent mothers. International Quarterly of Community Health Education. 1998;18(1):121–37. PubMed PMID: 107176677. Language: English. Entry Date: 19990401. Revision Date: 20150711. Publication Type: Journal Article.

31. Mitchell A, Gutmann-Gonzalez A, Brindis CD, Decker MJ. Contraceptive access experiences and perspectives of Mexican-origin youth: a binational qualitative study. Sex Reprod Health Matters. 2023;31(1):2216527. doi: 10.1080/26410397.2023.2216527. PubMed PMID: 37335382; PubMed Central PMCID: PMCPMC10281292.

32. Morales-Alemán MM, Ferreti G, Scarinci IC. “I don’t like being stereotyped, I decided I was never going back to the doctor”: Sexual healthcare access among young Latina women in Alabama. J Immigr Minor Health. 2020;22(4):645–52. doi: 10.1007/s10903-019-00932-3. PubMed PMID: 31535273; PubMed Central PMCID: PMCPMC7078038.

33. Norris AE, Ford K. Beliefs about condoms and accessibility of condom intentions in Hispanic and African American youth. Hisp J Behav Sci. 1992;14(3):373–82. Epub 1992/08/01. doi: 10.1177/07399863920143007. PubMed PMID: 12345005.

34. Wilson EK, Samandari G, Koo HP, Tucker C. Adolescent mothers’ postpartum contraceptive use: a qualitative study. Perspect Sex Reprod Health. 2011;43(4):230–7. Epub 2011/12/14. doi: 10.1363/4323011. PubMed PMID: 22151510.

35. Carvajal DN, Gioia D, Mudafort ER, Brown PB, Barnet B. How can Primary Care Physicians Best Support Contraceptive Decision Making? A Qualitative Study Exploring the Perspectives of Baltimore Latinas. Womens Health Issues. 2017;27(2):158–66. Epub 2016/11/09. doi: 10.1016/j.whi.2016.09.015. PubMed PMID: 27825590.

36. Kramer RD, Higgins JA, Godecker AL, Ehrenthal DB. Racial and ethnic differences in patterns of long-acting reversible contraceptive use in the United States, 2011-2015. Contraception. 2018;97(5):399–404. Epub 20180210. doi: 10.1016/j.contraception.2018.01.006. PubMed PMID: 29355492; PubMed Central PMCID: PMCPMC5965256.

37. Barral RL, Cartujano B, Perales J, Ramirez M, Cowden J, Trent M, et al. Knowledge, beliefs, and attitudes about contraception among rural Latino adolescents and young adults. The Journal of Rural Heal*th*2020. p. 38–47.

38. Guilamo-Ramos VB, A. S.: Benzekri, A.: Ruiz, Y.: Beltran, O. Misalignment of sexual and reproductive health priorities among older Latino adolescents and their mothers. Contraception. 2019;99(3):179–83. Epub 2018/11/25. doi: 10.1016/j.contraception.2018.11.011. PubMed PMID: 30471265.

39. Barral RL, Cartujano B, Cupertino AP, Trent M. “Birth control is like having an abortion: Attitudes, beliefs and knowledge about sex and contraception among Latino youth in rural kansas”. Journal of Adolescent Health. 2015;56(2):S39. doi: 10.1016/j.jadohealth.2014.10.079.

40. Patel PR, Lee J, Hirth J, Berenson AB, Smith PB. Changes in the Use of Contraception at First Intercourse: A Comparison of the National Survey of Family Growth 1995 and 2006-2010 Databases. J Womens Health (Larchmt). 2016;25(8):777–83. Epub 2016/02/27. doi: 10.1089/jwh.2015.5513. PubMed PMID: 26919078; PubMed Central PMCID: PMCPMC4982959.

41. American Academy of Pediatrics. The importance of access to comprehensive sex education. February 15, 2024.

